# SARS-CoV-2 vaccine and increased myocarditis mortality risk: A population based comparative study in Japan

**DOI:** 10.1101/2022.10.13.22281036

**Authors:** Sintaroo Watanabe, Rokuro Hama

**Author notes:** **Correspondence to: Rokuro Hama,** Non-profit Organization “Japan Institute of Pharmacovigilance (Med Check)”, **Email:**. **CONTRIBUTIONS:** SW proposed to conduct the study, RH and SW designed the study. Both authors independently converted the pdf data disclosed by the Japanese government into excel files and confirmed data matched. Then data collection, classification, and analyses were done first by the SW and later confirmed by RH, with any discrepancies being discussed and decided by both parties. SW and RH contributed equally to this study and reviewed the final manuscript. **COMPETING INTERESTS:** All authors have completed the ICMJE uniform disclose form available at https://www.icmje.org/disclosure-of-interest/, RH wrote a book entitled “Drugs to avoid, and infectious diseases including COVID-19” published on 1 December 2020. **ETHICAL APPROVAL** This study was conducted based only on data disclosed by the Japanese government. Japanese law and guidelines do not require an ethical approval for such research. **DISCLAIMER** The content is the personal view of the author and is not related to the official view of the authors’ organization.

## Abstract

**Objective:** To investigate the association between SARS-CoV-2 vaccination and myocarditis death

**Design:** Population based comparative mortality study

**Setting:** Japan

**Participants:** Vaccinated population was 99 834 543 individuals aged 12 years and older who have been received SARS-CoV-2 vaccine once or twice by 14 February 2022. Reference population was defined persons aged 10 years and older from 2017 to 2019.

**Main outcome measures:** The primary outcome was myocarditis death, defined as the case with “myocarditis” for primary cause of death and with onset 28 days or less after vaccination disclosed on 11 November 2022. Myocarditis mortality rate ratio (MMRR) of the SARS-CoV-2 vaccinated to the reference population by 10-year age group and standardised mortality ratio (SMR) were calculated. Mortality odds ratios (MORs) by 10-year age group were also calculated for supplementary analysis. Healthy vaccine effect-adjusted MMRRs (adMMRRs) or adjusted SMR (adSMR) were calculated by dividing MMRRs or SMR by 0.24 respectively.

**Results:** Number of myocarditis death which met the inclusion criteria were 32 cases. MMRR (95% confidence interval) was 4.03 (0.77 to 13.60) in 20s, 7.80 (2.85 to 18.56) in 30s, respectively. SMR of myocarditis was 1.69 (1.18 to 2.42) for overall vaccinated population, 1.35 (0.84 to 2.55) for those 60 years or older. Estimated adMMRRs and adSMR were about 4 times higher than the MMRRs and SMR. Pooled MOR for myocarditis were 148.49 (89.18 to 247.25).

**Conclusion:** SARS-CoV-2 vaccination was associated with higher risk of myocarditis death, not only in young adults but also in all age groups including the elderly. Considering healthy vaccinee effect, the risk may be 4 times or higher than the apparent risk of myocarditis death. Underreporting should also be considered. Based on this study, risk of myocarditis following SARS-CoV-2 vaccination may be more serious than that reported previously.

**SUMMARY BOXES:** *ALREADY KNOWN ON THIS TOPIC:* There are many epidemiological studies showing increased myocarditis incidence after SARS-CoV-2 vaccination. There are also some case reports of fulminant myocarditis after receiving SARS-CoV-2 vaccine. However, no epidemiological studies focusing the association between vaccination and myocarditis death.

*WHAT THIS STUDY ADDS:* Myocarditis mortality rate ratios (MMRRs) and their 95% confidence intervals (95% CIs) after receiving SARS-CoV-2 vaccine compared with that in the reference population (previous 3 years) were higher not only in young adults (highest in the 30s with MMRR of 7.80) but also in the elderly. Standardised mortality ratio (SMR) for myocarditis was 1.35 (0.84 to 2.55) for those 60 years or older and 1.69 (1.18 to 2.42) in overall age. The risk of myocarditis mortality in the SARS-CoV-2 vaccinated population may be 4 times or higher than the apparent MMRRs considering healthy vaccinee effect. Unreported post-vaccination deaths should also be considered as suggested by the extremely high myocarditis mortality odds ratio (148.49; 89.18 to 247.25).

## INTRODUCTION

Amongst the several safety concerns of SARS-CoV-2 vaccination, myocarditis is one of the most important adverse reactions which package insert warn as “Post marketing data demonstrate increased risks of myocarditis and pericarditis”.^1 2^ After a notification of a possible link between SARS-CoV-2 vaccination and myocarditis by the US Centers for Disease Control and Prevention (CDC),^3^ a number of case reports have been published ^4^ and several fatal cases have also been published.^5-12^ In Japan, a 27-year-old professional athlete with no history of symptomatic illness except orthopedic problem was rushed to hospital with cardiac arrest on day eight of the first dose of mRNA-1273 (Moderna) vaccine and subsequently died, with autopsy results revealing myocarditis.^12^ A non-comparative epidemiological study indicated that the highest incidence of myocarditis was reported in male patients between the ages of 16 and 29 years.^13^ Comparative epidemiological studies showed that SARS-CoV-2 vaccination is associated with increased risk of myocarditis especially in adolescent and young adults without exception.^14-18^ However, they reported that myocarditis after vaccination was mild^14 16^ and did not focus on the fatal cases.^14-18^ The package inserts of SARS-CoV-2 vaccine do not mention the possibility of death from myocarditis after the vaccination.^1 2^ To our best knowledge, no epidemiological studies have conducted to investigate the association of increased risk of SARS-CoV-2 vaccine on myocarditis death.

The primary objective of this study is to investigate association between SARS-CoV-2 vaccine and myocarditis deaths comparing mortality rate with general population, then to provide a new discussion on the healthy vaccinee effect in using SARS-CoV-2 vaccine, especially on death. In addition, all-cause death after vaccination was also investigated and under-reporting of deaths was discussed.

## METHODS

This study compared myocarditis mortality rate in the SARS-CoV-2 vaccinated with that in the general population in Japan. The study was based on the materials and the vital statistics disclosed by the Japanese government.

### Data sources and case definition

#### 1. Vaccinated population

The vaccinated population was defined as those who had received the first or the second doses of SARS-CoV-2 vaccine since the start of the vaccination program (17 February 2021) until 14 February 2022. The number of persons received vaccine by number of doses was disclosed by the Japanese Cabinet Office by 10-year age group without background information for sex, vaccine type and others.^19^ According to this information, 99 834 543 persons received the first dose and 99 117 143 persons received the second dose. Of the Japanese population aged 12 years or older, 89.6% received at least one dose of SARS-CoV-2 vaccine.

The number of vaccinated populations by number of doses, by vaccine type for all age groups combined was also disclosed.^20^ This information was used to estimate the interval between the first and the second dose. The discrepancy between the total number of vaccinees by age group and the total number of vaccinees by vaccine type may be due to the presence of vaccination cases of unknown age group at the time of reporting.

#### 2. Death cases after receiving SARS-CoV-2 vaccine

Data for the death cases after receiving SARS-CoV-2 vaccine were based on “the summary list of death cases after SARS-CoV-2 vaccination” as pdf files that were disclosed by the panels of experts on vaccination and adverse reactions under Japan’s Ministry of Health, Labour and Welfare (MHLW) at 11 November 2022.^21-23^ The list include age, sex, number of vaccination, last vaccinated date, comorbidities, histories leading to death, cause of death diagnosed by physician, corresponding MedDRA terms, tests for the base of diagnosis, causal assessment by physician, assessment by the experts (Fig S1). Both authors independently converted the pdf data into excel files and confirmed data matched. Total number of death cases reported by 9 October and disclosed on 11 November was 1 897 (Fig 1).

**Fig 1.**
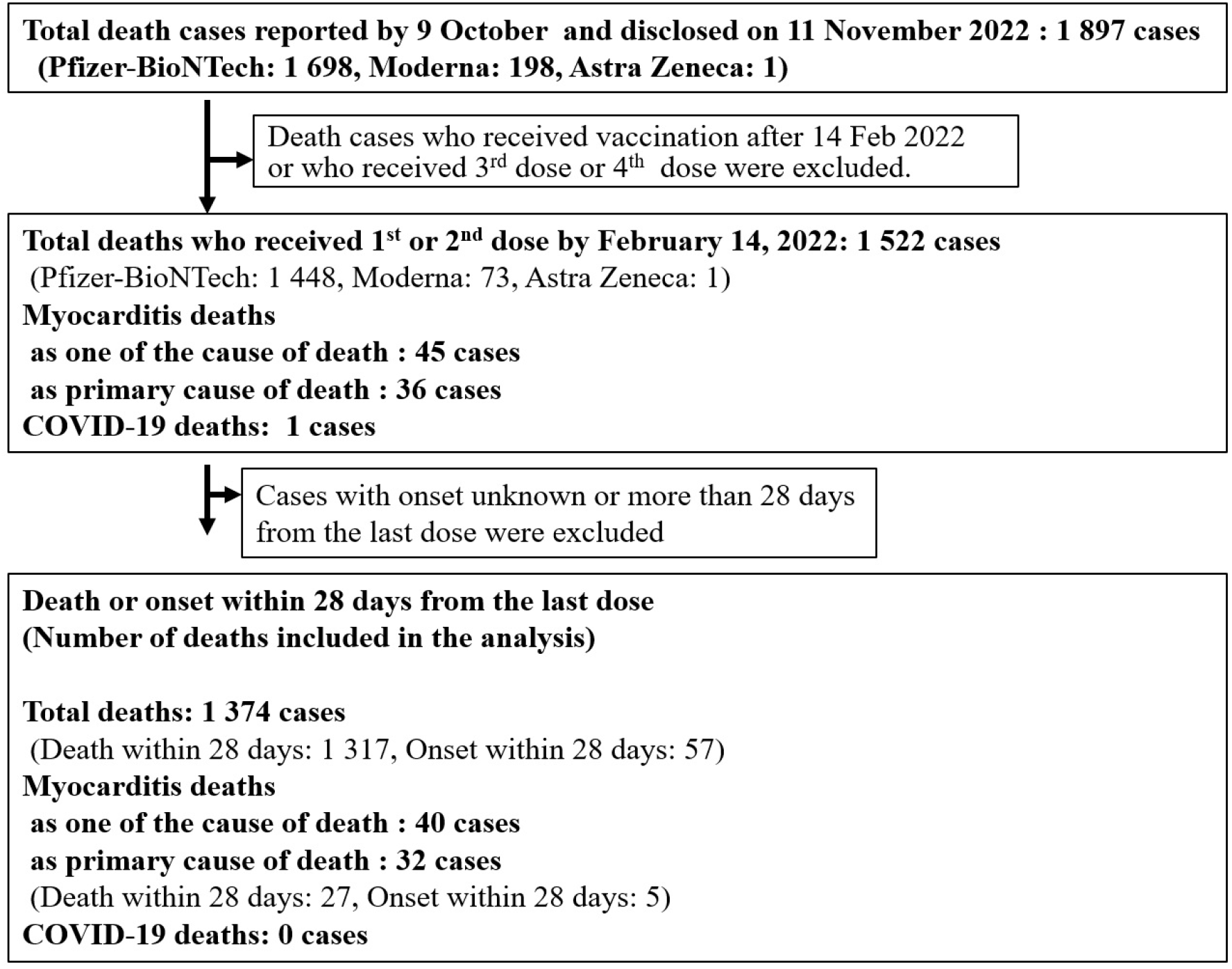
Included death cases for analysis based on the list disclosed on 11 November 2022.

In Japan, doctors are required to report serious adverse reactions to vaccine in general including death within 28 days if they suspected an association with vaccination.^24^ For SARS-CoV-2 vaccine, those occurred during the period considered by a physician as highly relevant to vaccination were required to report at the beginning of the vaccination program.^24^ Subsequently, physicians were required active consideration of reporting myocarditis, pericarditis and thrombosis occurring within 28 days after vaccination if they suspected an association with vaccination. ^25^ Therefore, we defined the death cases for comparison of mortality rate as those in which the onset (start of signs and symptoms leading to death) was known as occurred within 28 days after the last dose of SARS-CoV-2 vaccine (reasons for inclusion of onset within 28 days are explained later in detail). We restricted the vaccinee who received one or two dose and excluded those with third or fourth dose to avoid further healthy vaccinee effect.^26 27^ Number of included death cases was 1 374 in total (Fig 1).

#### 3. Myocarditis death after SARS-CoV-2 vaccination

“Myocarditis death case” after vaccination was defined as the cases in which “myocarditis” was described in the cause of death column of the above summary list, regardless of diagnostic method. When more than one cause of death was listed, we extracted cases where “myocarditis” was listed as the primary cause of death. They were classified as death from I40 (acute myocarditis) under the 10th revision of the International Statistical Classification of Diseases and Related Health Problems (ICD-10).

On the 11 November 2022 list, number of included myocarditis death reports was 32, of which 22 were on the 18 February 2022 list and 10 new deaths were added since then.

Diagnostic bases for myocarditis are classified according to the information on the summary list of death cases disclosed on 11 November 2022 as follows: 1. Autopsy and/or myocardial biopsy, 2. Elevated Troponin with blood test, 3. Other blood test and/or (ECG and/or UCG), 4. Only symptoms. Diagnostic basis for the lower number does not include the diagnostic basis for the higher number.

#### 4. Calculation of person-years of exposure for vaccinated population

Average days from first to second dose are different among three products: 21 days for BNT162b, ^1^ 28 days for mRNA-1273, ^2^ and 40 days for ChAdOx1 nCoV-19.^28 29^ The number of second dose were 84 023 380 for BNT162b, 16 090 036 for mRNA-1273, and 58 300 for ChAdOx1 nCoV-19 (Table S1). However, the number of doses by age group and by vaccine type was not disclosed. Therefore, weighted average days from first to second dose for overall ages was estimated as 22.14 days in overall ages (Table S2).

Based on the above information, the person-years of observation for the first dose and the second dose were calculated as follows: D = (A ×22.14+B ×5.86)/365, E =C×28/365, F= D + E. where 22.14 is the weighted average days from the first dose to the second dose and 5.86 is 28 days of observation period - 22.12, A is the number of vaccinees who received the first dose, B is that with only the first dose, C is that with the second dose, D is the person-years for the first dose, E is for the second dose and F is total person-years.

#### 5. Reference population and death

We chose the general Japanese population during the period from 2017 through 2019 in the pre-COVID-19 pandemic era as reference population and death for comparison based on the vital statistics in 2017, 2018 and 2019.

Myocarditis mortality rates in the reference population were calculated from the total number of myocarditis deaths by total population by 10-year age groups in three years from 2017 to 2019.^30-35^

### Outcome measures and statistical analyses

Primary outcome measure was myocarditis mortality rate. We compared the observed mortality rate of myocarditis with the expected mortality rate using data for reference population. Myocarditis mortality rate ratios (MMRRs) and their 95% confidence intervals (95%CIs) by 10-year age group were calculated.

Sensitivity analysis was performed as follows:

1. Standardised mortality ratio (SMR) for overall age stratified by 10-year age groups.
2. SMRs for 3 age groups (12-39, 40-59 and 60 or older) stratified by 10-year age groups.
3. SMR by the vaccination status after the first or second dose.
4. SMR for all-cause death.
5. Mortality odds ratio (MOR) by 10-year age group for myocarditis applying reporting odds ratio (ROR).^36^
6. MMRRs and SMR adjusted by the healthy vaccinee effect were estimated: Healthy vaccinee effect by SARS-CoV-2 vaccination expressed as the rate ratio of mortality rate in vaccinated to that in the reference general population was shown approximately as 0.10 to 0.24 (95% CI were not given). ^26 27^ An approximate healthy vaccinee effect-adjusted MMRRs (adMMRRs) or adjusted SMR (adSMR) were yielded by dividing MMRRs or SMR by 0.10 to 0.24 respectively without 95% CI, by applying the prior event rate ratio (PERR) adjustment method for “unmeasured confounding” in observational studies.^37-39^

The valid reasons for comparing myocarditis mortality rate of vaccinee with onset within 28 days from the last dose and of the reference population are based on the following. In the reference population, death cases in which signs and symptoms of myocarditis leading to death develop during the observation period (From the beginning through the end of the year) and die after the observation period are not included in the mortality calculation, whereas cases that develop before the observation period and die during the observation period are included in the mortality calculation (Fig S2a).

In the post-vaccination population, on the other hand, a case in which signs and symptoms of myocarditis leading to death had already developed before vaccination would have never been reported as a death case possibly or probably associated with vaccine and is never included in the mortality analysis. Hence, cases in which signs and symptoms of myocarditis leading to death develop during the observation period should be included in the mortality analysis among vaccinated population for fair comparison with the reference population (Fig S2b).

Using the same method, number of all-cause deaths in the post-vaccinated population was calculated and compared with the expected number of deaths from all-cause.

All statistical analyses were performed with Stats Direct (Version 3.3.5). The significance level was set at *P*<0.05. Statistical multiplicity was not tested because this is rather an exploratory study but not a hypothesis confirming study.

Ethical approval was not obtained for this study because it was based on the disclosed data and is not required to obtain ethics approval under Japanese law and guidelines.

### Patient and Public involvement

Patients and public were not involved in the design, analyses, in this study because the research agenda was urgent.

## RESULTS

The number of included death cases for analysis was 1 374 including 32 myocarditis death and their characteristics are shown in Table 1.

**Table 1.** Characteristics of included death cases after receiving SARS-CoV-2 vaccine ^*a^.

|  | All-cause death<br>N (%) | Myocarditis death<br>N (%) |
| --- | --- | --- |
| **Total** | 1 374 (100.0) | 32 (100.0) |
| **Age (years)** |  |  |
| 12-19 | 7 (0.5) | 0 (0.0) |
| 20-29 | 31 (0.3) | 3 (3.3) |
| 30-39 | 33 (1.5) | 6 (6.6) |
| 40-49 | 54 (0.9) | 3 (3.3) |
| 50-59 | 91 (2.8) | 0 (0.0) |
| 60-69 | 124 (2.2) | 3 (3.3) |
| 70-79 | 302 (0.0) | 6 (6.6) |
| 80-89 | 453 (5.3) | 7 (7.7) |
| ≥ 90 | 265 (2.4) | 2 (2.2) |
| Unknown | 2 (0.1) | 0 (0.0) |
| **Sex** |  |  |
| Male | 750 (5.9) | 21 (65.6) |
| Female | 607 (6.6) | 11 (34.4) |
| Unknown | 5 (1.4) | 0 (0.0) |
| **Vaccine type** |  |  |
| BNT162b2 (Pfizer-BioNTech) | 1 298 (11.3) | 26 (81.3) |
| mRNA-1273 (Moderna) | 63 (1.7) | 6 (18.8) |
| ChAdOx1 nCoV-19 (Astra Zeneca) | 1 (0.1) | 0 (0.0) |
| **Number of doses of vaccination** |  |  |
| 1 | 753 (5.2) | 13 (40.6) |
| 2 | 556 (7.0) | 18 (56.3) |
| Unknown <sup>*b</sup> | 53 (0.9) | 1 (3.1) |

<sup>\*a</sup>: Onset of symptoms leading to death occurred within 28 days after receiving the last dose of vaccine.
<sup>\*b</sup>: Dose was surely first or second but not known which dose.
<sup>\*c</sup>: Diagnostic basis for the lower number does not include the diagnostic basis for the higher number.
<sup>\*d</sup>: Possibility could not be ruled out that "Other blood tests" include elevated troponin.

**Table 1(continued)| Characteristics of included death cases after receiving SARS-CoV-2 vaccine <sup>\*a</sup>**
|  | All-cause death<br>N (%) | Myocarditis death<br>N (%) |
| --- | --- | --- |
| **Time from vaccination to onset of symptom** |  |  |
| 0 to 7 days | 979 (11.1) | 16 (50.0) |
| 8 to 14 days | 213 (1.6) | 7 (21.9) |
| 15 to 28 days | 170 (0.4) | 9 (28.1) |
| **Diagnostic basis for myocarditis deaths <sup>\*c</sup>** |  |  |
| 1.Autopsy and /or myocardial biopsy |  | 16 (50.0) |
| 2.Elevated Troponin with blood test |  | 3 (9.4) |
| 3.Other blood test <sup>*d</sup> and/or (ECG and/or UCG) |  | 10 (31.3) |
| 4.Only symptoms |  | 1 (3.1) |
| 5.Unknown |  | 2 (6.3) |

\*a: Onset of symptoms leading to death occurred within 28 days after receiving the last dose of vaccine.
\*b: Dose was surely first or second but not known which dose.
\*c: Diagnostic basis for the lower number does not include the diagnostic basis for the higher number.
\*d: Possibility could not be ruled out that "Other blood tests" include elevated troponin.

Persons who died from myocarditis death were younger, used more mRNA-1273 and occurred more after the second dose than those from other causes after SARS-CoV-2 vaccination. Myocarditis was diagnosed by autopsy and/or myocardial biopsy in half cases.

Table 2 shows the number of vaccinated persons by age and the person-years of observation calculated from them. Table 3 shows the population, causes of death, and their crude mortality rates for 2017-2019, used as reference. Details for each year are shown in the Table S3.

**Table 2.** Number of persons who received vaccine by dose and person-years of exposure by age.

| Age<br>(years) | Number of persons who received |  |  | Person-years |  |  |
| --- | --- | --- | --- | --- | --- | --- |
|  | A. First<br>dose, N | B. Only first<br>dose, N | C. Second<br>dose, N | D. First<br>dose, N | E. Second<br>dose, N | F. Total<br>N (%) |
| 12-19 | 6 881 439 | 147 798 | 6 733 641 | 419 700 | 516 553 | 936 253 (6.8) |
| 20-29 | 10 272 284 | 141 353 | 10 130 931 | 625 234 | 777 167 | 1 402 401 (10.3) |
| 30-39 | 11 526 243 | 120 385 | 11 405 858 | 700 943 | 874 970 | 1 575 913 (11.5) |
| 40-49 | 15 392 743 | 99 856 | 15 292 887 | 935 098 | 1 173 153 | 2 108 250 (15.4) |
| 50-59 | 15 331 940 | 65 721 | 15 266 219 | 930 862 | 1 171 107 | 2 101 969 (15.4) |
| 60-69 | 13 926 130 | 38 967 | 13 887 163 | 845 176 | 1 065 317 | 1 910 493 (14.0) |
| 70-79 | 15 300 344 | 43 875 | 15 256 469 | 928 595 | 1 170 359 | 2 098 954 (15.4) |
| 80-89 | 8 875 050 | 41 716 | 8 833 334 | 538 898 | 677 626 | 1 216 523 (8.9) |
| ≥ 90 | 2 328 370 | 17 729 | 2 310 641 | 141 489 | 177 255 | 318 744 (2.3) |
| Total | 99 834 543 | 717 400 | 99 117 143 | 6 065 995 | 7 603 507 | 13 669 500 (100) |

$D = (A \times 22.14 + B \times 5.86) / 365$ where 22.14 was the weighted average days from first dose to second dose and 5.86 is 28 days of observation period - 22.12. $E = C \times 28 / 365$

**Table 3.** Population, cause of death and mortality rate in the reference population by age.

| Age<br>(years) | Population <sup>a</sup> | Number of deaths by cause <sup>a</sup> |  | Crude mortality rate (/100,000) |  |
| --- | --- | --- | --- | --- | --- |
|  | (2017~2019)<br>N (%) | All-cause<br>N (%) | Myocarditis<br>N (%) | All-cause | Myocarditis |
| 10-19 | 33 897 000 (9.7) | 4 807 (0.1) | 16 (3.5) | 14 | 0.05 |
| 20-29 | 37 697 000 (10.8) | 12 621 (0.3) | 20 (4.3) | 33 | 0.05 |
| 30-39 | 43 927 000 (12.6) | 23 183 (0.6) | 25 (5.4) | 53 | 0.06 |
| 40-49 | 56 179 000 (16.1) | 67 014 (1.6) | 48 (10.4) | 119 | 0.09 |
| 50-59 | 48 037 000 (13.8) | 140 399 (1.6) | 54 (11.7) | 292 | 0.11 |
| 60-69 | 50 918 000 (14.6) | 388 709 (3.4) | 92 (19.9) | 763 | 0.18 |
| 70-79 | 45 582 000 (13.1) | 819 510 (9.5) | 101 (21.8) | 1 798 | 0.22 |
| 80-89 | 26 491 000 (7.6) | 1 499 479 (20.1) | 84 (18.1) | 5 660 | 0.32 |
| ≥ 90 | 6 545 000 (1.9) | 1 118 796 (36.8) | 23 (5.0) | 17 094 | 0.35 |
| Total | 349 273 000 (100) | 4 074 518 (100) | 463 (100) | 1 167 | 0.13 |

<sup>a</sup>a: Population and number of deaths were all total of 3 years

### Myocarditis mortality in SARS-CoV-2 vaccinated population compared with reference population

MMRRs and their (95%CI) after receiving SARS-CoV-2 vaccine were 4.03 (0.77 to 13.60) in 20s, 7.80 (2.85 to 18.56) in 30s. Except 10s and 50s and 60s, each point estimates of the MMRR exceeded 1.00. Crude MMRR was 1.77 (1.19 to 2.53) for overall vaccinated population, 1.62 (0.85 to 2.79) after the first dose and 1.79 (1.05 to 2.85) after the second dose.

SMR of myocarditis was 1.69 (1.18 to 2.42) for overall vaccinated population, 1.53(0.88 to 2.66) after the first dose and 1.72(1.07 to 2.76) after the second dose. Details were shown in Fig 2 and Table 4. SMR of myocarditis in those aged 12-39, 40-59 and the elderly overall (60 years or older) was 4.52 (2.32 to 8.82), 1.01 (0.37 to 2.74) and 1.35 (0.84 to 2.17), respectively.

**Table 4.**
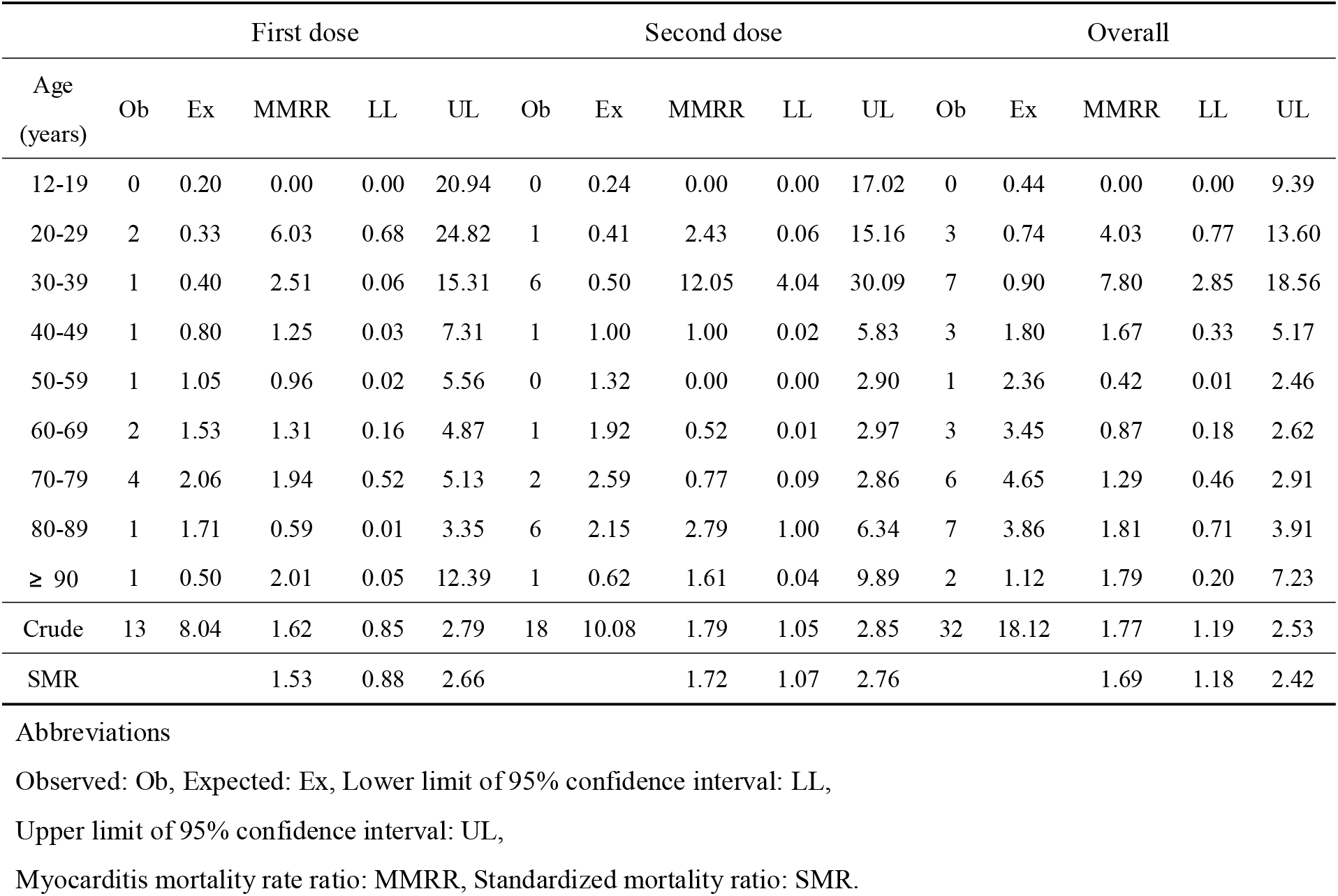
Myocarditis mortality rate ratios (MMRRs) by age by dose of vaccination.

**Fig 2.**
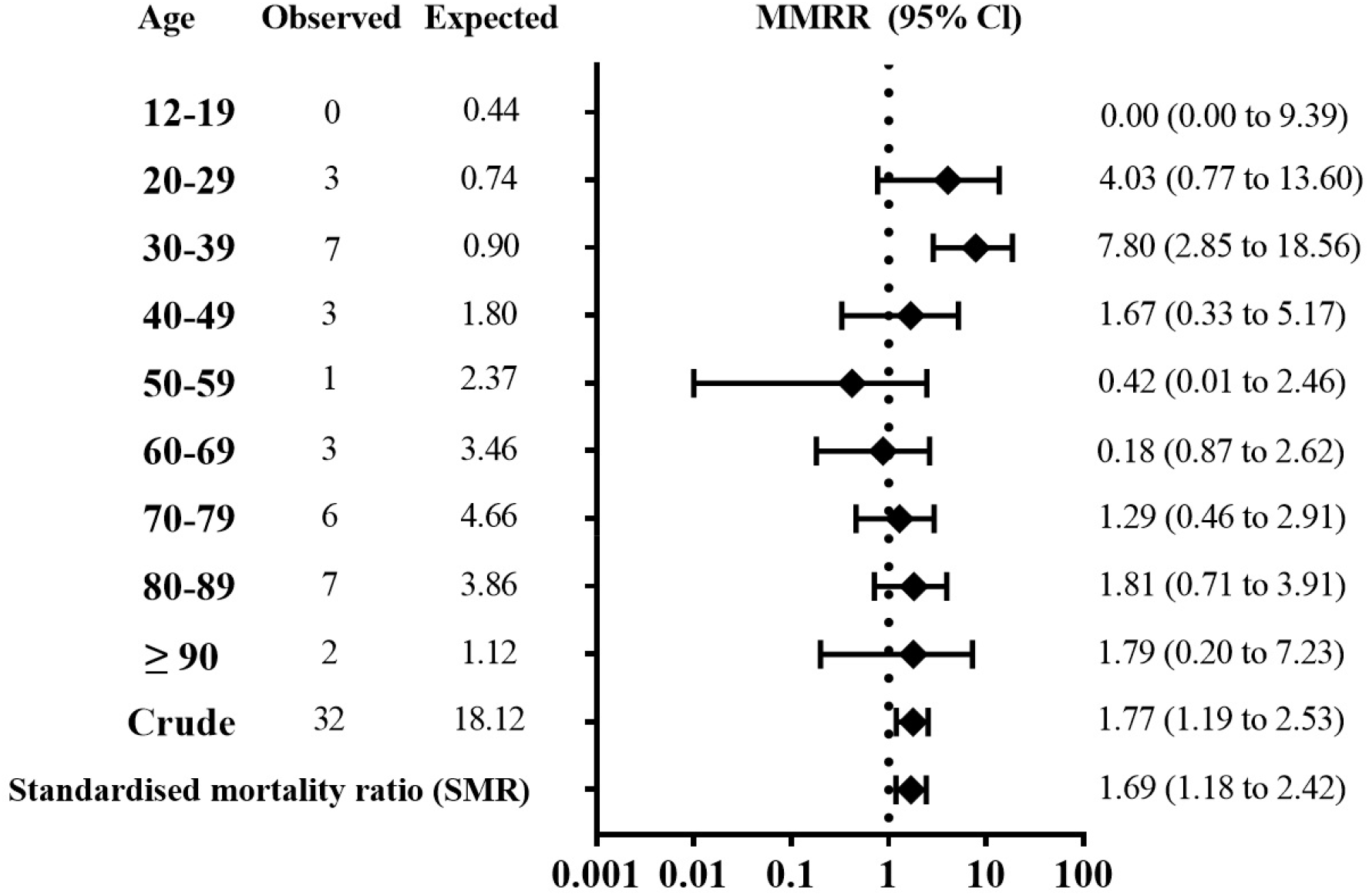
Myocarditis mortality rate ratios (MMRRs) by age and Standardised mortality ratio (SMR).

### All-cause mortality in SARS-CoV-2 vaccinated population

The apparent rate ratios for all-cause death after receiving SARS-CoV-2 vaccine were shown in Table S4. The apparent SMR for all-cause mortality was 0.01.

### Mortality odds ratio for myocarditis

Age-stratified and pooled MORs for myocarditis death after receiving SARS-CoV-2 vaccine were shown in Table S5. Except 10s in which reported myocarditis death were zero, point estimates MORs for myocarditis were above 20. The crude MOR was 209.82 (146.06 to 301.41), and the pooled MOR (95% CI) for myocarditis death was 148.49 (89.18 to 247.25).

### Healthy vaccinee effect adjusted MMRRs

Estimated point adMMRRs were approximately 17 to 40 in 20s, 33 to 80 in 30s, and adSMR was 7 to 17 in overall vaccinated population (95%CI were not calculated).

## DISCUSSION

### Principal findings

Using the disclosed data by the Japanese government, we observed increased myocarditis mortality rate ratio in the SARS-CoV-2 vaccinated population compared with general population during three years pre-COVID-19 pandemic era, especially in young adults (MMRR: 7.80 in 30s). However, not only in young adults, but also in the elderly and overall vaccinated, increased risk of myocarditis death is associated with vaccination even without consideration of healthy vaccinee effect. The pooled MOR for myocarditis death was as high as 148.49. Very rough estimation of healthy vaccinee effect adjusted MMRRs showed as high as 33 to 80 in 30s and about 7 to 17 for adSMR which were closer to pooled MOR. Increased risk was higher after the second dose than the first dose as shown in previous epidemiologic studies focusing on hospitalised myocarditis patients.^14-18^

### Strengths of this study

This study has several strengths. First this is the first epidemiological study that show the increased risk of myocarditis mortality after the SARS-CoV-2 vaccination. Previous epidemiological studies which reported the increased risk of myocarditis did not report the increased death from myocarditis ^14-18^ with emphasis that most cases were mild,^14 16^ that deaths were rare with no deaths of persons younger than 40 years ^17^ and that one person died with fulminant myocarditis in the nationwide Israeli study.^14^

Patone et al. ^40^ conducted a self-controlled case series study focusing on incidence but not on mortality from myocarditis following vaccination. However, using their data ^40^ and the statistics of England, ^41 42^ we found similar evidence of increased risk of myocarditis mortality in SARS-CoV-2 vaccinated population as shown in our study.

In their paper, ^40^ there was the number of persons received SARS-CoV-2 vaccine in the Table 1; there was the number of myocarditis deaths occurred within 1-28 days after receiving vaccine in the Supplementary Table 1. ^40^ Sixty-one myocarditis deaths have been observed during an overall observation period of 54.24 (100 000 person-years) for the entire vaccinated population aged 16 years and older; hence myocarditis mortality rate was 1.12 per100 000 person-years for the post-vaccinated population. In the three-year period from 2017 to 2019, the observation period of England aged 15 years and older was 1 375 (100 000 person-years), ^41^ during which 552 deaths for myocarditis ^42^ were observed (myocarditis classification in England is different from Japanese classification). The myocarditis mortality rate from 2017 to 2019 in England was therefore 0.40 per 100 000 person-years. Based on these data, crude MMRR in England would be 2.80 (2.11 to 3.65) if calculating by the same method of ours.

Other study with the most vaccinated population was a study of approximately 19 million persons in four Nordic countries.^17^ Although some parts of the discussion and Supplementary Content in their article referred to deaths, their study was not primarily aimed at investigating post-vaccination myocarditis deaths. In their study, about 6 million of those vaccinated were under 39 years of age, about one-fifth of those in this study; therefore, it may be difficult to detect an increase in myocarditis deaths among younger subjects.

Second, we revealed increase risk of myocarditis death in all age groups even without consideration of healthy vaccinee effect.

Third, we pointed out that if healthy vaccinee effect was considered, the risk of SARS-CoV-2 vaccine on myocarditis death may be much higher, with rate ratio up to 33 to 80 in 30s.

Fourth in addition, we showed that pooled MOR is extremely high although it is assumed that reports of cases with myocarditis after SARS-CoV-2 vaccine may be enhanced because it was widely reported in the media.

Fifth these results indicated that myocarditis mortality is increased in vaccinated persons and provide important insights into the consideration of benefits and harms of the SARS-CoV-2 vaccine.

### Limitations of this study

This study has several limitations. First, diagnosis of myocarditis death after SARS-CoV-2 vaccine was based on the physician’s diagnosis and was not based on exactly the newly proposed Brighton’s case definition.^43^ However, 50 % of myocarditis cases were diagnosed by autopsy and/or myocardial biopsy. They are exactly the level 1 myocarditis (definite case) by the Brighton’s case definition. Including above definite cases, almost 90% of cases were diagnosed at least blood test and/or ECG and/or UCG. Cases based on symptoms only or unknown methods were 11%. Moreover, diagnostic base of the myocarditis death in the reference population from 2017 to 2019 was not known and was also based on physician’s diagnosis. Previous studies ^14-18^ supported a high incidence of post-SARS-CoV-2 vaccination myocarditis, making it difficult to consider that diagnostic accuracy is a factor in overestimating myocarditis mortality after vaccination. As described in the strengths of this study, the risk of myocarditis mortality following vaccination in the 2017-2019 population in England may be 2.80 (2.11 to 3.65) as a crude MMRR. In contrast, in this study, the crude MMRR was 1.77 (1.19 to 2.53) which is similar to the SMR and even lower than that in England.

Second, because myocarditis after SARS-CoV-2 vaccine received media attention, it is likely that physicians paid more attention and reported more. It might be one of the reasons for extremely high pooled MOR for myocarditis death. However, this may be resulted from underreporting of death from other causes, because not all deaths after SARS-CoV-2 vaccine were reported. There is no obligation to report all post-vaccination deaths in Japan, therefore only those cases where a physician suspected association to vaccination are reported and disclose by the MHLW. In the United States (US), 610 million doses of the SARS-CoV-2 vaccine had been administered until 31 August 2022, and about 16 000 post-vaccination deaths had been reported, ^44^ while in Japan, only about 1 500 post SARS-CoV-2 vaccination deaths had been disclosed against 200 million doses of the vaccine until 14 February 2022. If post-vaccination deaths were reported on a par with the US, this could be about 3.5 times higher. The apparent SMR for all-cause deaths were as low as 0.01. It is far lower than those estimated by other data. For example, randomized control trials have shown that the SARS-CoV-2 vaccine was not effective in reducing all-cause death.^45 46^ The results of analysis ^26 47^ using data of UK statistics ^48^ showed that age adjusted non-COVID-19 mortality rate in the ever vaccinated compared with that in the general population was estimated at 0.61 in January 2021. These results indicate that there was substantial underreporting of SARS-CoV-2 post-vaccination deaths. Mevorach revealed that the risk of myocarditis morbidity after vaccination was highest among second-time vaccinators aged 16-19 years.^14^ On the other hand, Japan’s MHLW database had not yet listed any myocarditis deaths between the ages of 12 and 19 years. Subsequently, vaccination of people aged 5-11 years began in Japan, and as of 11 November 2022, one person had died from myocarditis.^49^

Third, SMR was only adjusted for age, MMRRs and SMR were not adjusted for sex and for other factors such as calendar period, health care worker status, nursing home resident, and comorbidities like Husby et al ^16^ and/or Karlstad et al. ^17^ Mevorach et al, who compared the incidence of myocarditis between the three-year period 2017-2019 and the post-vaccination population in Israel, also stated that they had not adjusted for factors other than age and sex as a limitation of their study. However, to our best knowledge, obvious confounding factors ^50^ were not found that would overestimate myocarditis mortality following SARS-CoV-2 vaccination in this study. A cohort study by Karlstad et al.^17^ investigated the incidence of myocarditis in vaccinated and unvaccinated groups; we could compare of crude incidence rate ratios (IRRs) and adjusted IRRs using the data (Table 2 and eFigure2 in their paper). Even if the model with the lowest adjusted IRR was adopted (Adjusted by age group, sex, previous SARS-CoV-2 infection, healthcare worker, nursing home resident, comorbidity, and calendar time), the adjusted IRR was about 0.77 to 0.98 times compared to the crude IRR (2 to 23 % decrease from crude IRR). It does not extend to the impact of the underestimation of post-vaccination myocarditis mortality by healthy vaccine effect (underestimation by 0.10 to 0.24). Hence, the MMRRs and SMR may not be overestimated by this limitation.

Forth, we could not compare relative risk among products, BNT162b2 (Pfizer-Biotech) or mRNA-1273 (Moderna), because precise data for number of persons who received each product by age group until the cut off day (14 February 2022) were not disclosed. However, according to the analysis by National Institute of infectious diseases, ^51^ proportion of reports for myocarditis among male 10s and 20s who received second dose of mRNA-1273 was 102.1 and 47.2 per million persons respectively, while 15.4 and 10.0 per million persons for second dose of BNT162b2. If the distribution of number of persons who received each product by age group at the cut off day (14 February 2022) were the same as reported on 3 December 2021, mortality rate ratio from myocarditis among those who were younger than 40 years and received mRNA-1273 compared with those who received BNT162b2 show no significant increase: 3.12 (95%CI: 0.84 to 11.63, *P*=0.073), while it was not significant among those 40 years or older.

Fifth, we have no direct evidence on healthy vaccinee effect of SARS-CoV-2 vaccine in Japan. However, there are more than one evidence that indicates healthy vaccinee effect of SARS-CoV-2 vaccine in the world. One ^26 47^ is the results of analysis using data from UK statistics ^52^ and the other ^27 44^ is the results of analysis using the published data in the peer review journal.^53 54^

According to the former analysis, COVID-19-related mortality rate ratio (MRR) of those who died 21 days or more after the second dose to the unvaccinated was 0.02 at the beginning of the immunization program in UK (January 2021), while the non-COVID-19 MRR of those who died 21 days or more after the second dose to the unvaccinated was 0.11 (95%CI: 0.08 to 0.14) for January 2021 and 0.13 (0.10 to 0.17) for February 2021. These indicate that healthy vaccinee effect may work to lower the apparent risk of death from COVID-19 and may increase the apparent effectiveness of SARS-CoV-2 vaccine. Dividing the COVID-19-related MRR of 0.02 by the non-COVID-19 MRR of 0.11, yields a healthy vaccinee effect-adjusted COVID-19-related MRR of 0.18 (95%CI: 0.09 to 0.37). This may be considered to be closer to the true COVID-19-related MRR.^26^

Under similar conditions to this study, the healthy vaccinee effects estimated using UK statistics are as follows. Healthy vaccinee effect by SARS-CoV-2 vaccination expressed as MRR in ever vaccinated to expected mortality rate for 2021 assuming that COVID-19 is not epidemic in England and Wales (932.1/100 000 person years) was estimated 0.61 in January 2021 and 0.10 to 0.24 at the day of vaccination, by applying the analysis results ^27^ using data from Israeli study, ^53^ as shown in the following another evidence.

The analysis results show that odds ratio (OR) of symptomatic COVID-19 on day 1 of vaccination was 0.40 (95%CI: 0.31 to 0.51) and ORs of hospitalisation, severe COVID-19 and death due to COVID-19 on day 1 are roughly estimated as 0.27, 0.18 and 0.13 respectively (95%CI was not calculated). ^27^ Vaccination can never work at the day of vaccination; these low risk of mortality and morbidity is highly probably derived from the fact that the vaccinated people were much healthier than the unvaccinated. This bias could not be adjusted by ordinary methods for matching by adjusting age, sex, sector, and residence, history of influenza vaccination, pregnancy and total number of coexisting risk factors that Dagan et al used. This bias is a sort of “unmeasured confounding” in observational studies that Tannen et al proposed to adjust by the prior event rate ratio (PERR). ^37-39^

Theoretical basis of healthy vaccinee effect was shown by Fine et al. ^55^ Jackson et al ^56^ reported that the relative risk of death, hospitalisation due to pneumonia and ischemic heart disease for influenza vaccinated persons compared with unvaccinated persons was 0.36 (95%CI: 0.30 to 0.44), 0.65 (0.53 to 0.80) and 0.92 (0.83 to 1.02) before influenza season respectively in the United States. They concluded that the reductions in risk before influenza season indicate preferential receipt of vaccine by relatively healthy seniors and adjustment for diagnosis code variables did not control for this bias^53^ just as in the Dagan’s study ^53^ and Tannen et al. ^37-39^

These results indicate that the more serious the disease, the lower the apparent risk of vaccination and are consistent with the results of analysis ^27^ on Dagan’s data.^53^

In fact, Husby et al ^16^ mentioned the fact that SARS-CoV-2 vaccines are rarely given to people with an acute or terminal illness as a likely explanation of low 28-day risk of cardiac arrest or death in their study. This explanation is exactly the “healthy vaccinee effect”.

Considering these, healthy vaccinee effect works in the direction of positive for vaccination (more effective and safer) universally on observational studies even if many variables were matched and/or adjusted by the ordinary methods used in most observational studies including propensity score matching. Hence, it may be rational to take the health vaccinee effect into account in the present study. Because death is a rare event, the extent of the healthy vaccinee effect on death which were estimated using previously disclosed data showed wide range (0.10 to 0.24). If the least healthy vaccinee effect (the highest MRR: 0.24) is used, risk of SARS-CoV-2 vaccine on myocarditis death is estimated about 4 times higher than those without adjustment.

Lastly, this study is rather an exploratory study but not a hypothesis confirming study. However, we found several strong associations especially in the age of 30s even without adjustment for healthy-vaccinee effect and that very high MMRR were estimated if they were adjusted for healthy-vaccinee effect. Moreover, we got very high age-stratified and pooled MOR for myocarditis death. Hence, we discuss the causal inference on the increased myocarditis mortality and SARS-CoV-2 vaccine use primarily according to the modified criteria of US Advisory Committee to the Surgeon General ^57^ (modified US Surgeon General criteria) with some supplementary discussion using viewpoints of causation by Hill ^58^ (Hill’s viewpoints) (Table S6). Because “specificity of association” both in US Surgeon General criteria and Hill’s viewpoints is an extreme type of “strength of association”, we included it into “strength of association” and classified into 4 criteria: (1) temporarily, (2) consistency, (3) strength and (4) coherence of association.

We found all 4 criteria were satisfied and we conclude that the association of high myocarditis mortality rate ratio after SARS-CoV-2 vaccination may be causal.

### Points to be clarified in future research

Post-vaccination death should be more precisely investigated, not only from myocarditis but also from other causes. These should be closely monitored by nation-wide investigation as done in England and Wales or in the country with larger population. In these investigations, “healthy vaccinee effect” must be taken into account.

### Conclusions and policy implications

Despite above limitations, this study revealed that SARS-CoV-2 vaccination was associated with higher mortality rate from myocarditis, especially in young adults compared with 2017 to 2019 population. But it also revealed that myocarditis death occurs in older persons. If healthy vaccinee effect is taken into account, the risk increases at least approximately 4 times more than the unadjusted mortality risk. In addition, underreporting deaths after receiving vaccine should be considered. Based on the results of this study, it is necessary to inform public about that the risk of serious myocarditis including death may be far more serious than the risk reported before and that it occurs not only in young persons but also in elderly.

## Data Availability

All data produced in the present study are available upon reasonable request to the authors.

