## Supplemental files for "SARS-CoV-2 vaccine and increased myocarditis mortality risk: A population based comparative study in Japan"

**Supplemental Materials**

**Contents**

**Supplemental Tables**


Table S1. Number of first and second dose by vaccine type until 14 February 2022.

Table S2. Calculation of the average SARS-CoV-2 vaccination interval between the first and second dose.

Table S3. Characteristics of all-cause and myocarditis death in reference population.

Table S4. Rate ratio for all-cause death by age by dose of vaccination.

Table S5. Mortality odds ratio for myocarditis death by age.

Table S6. Causal inference of increased risk of myocarditis mortality and SARS-CoV-2 vaccine use by applying modified US Surgeon General's criteria of causality referencing Hill's viewpoints.


**Supplemental Figures**


Figure S1. Examples of description in the Summary List of Deaths Following SARS-CoV-2 vaccination used by Japan's Ministry of Health, Labour and Welfare's Expert Committee on vaccination and adverse reactions.

Figure S2a. Method for counting deaths in the reference population.

Figure S2b. Method for counting deaths in the vaccinated population.

**Table S1. Number of first and second dose by vaccine type until 14 February 2022.**

| | Number of first dose | Number of second dose | Total |
|---|---|---|---|
| BNT162b2 (Pfizer-BioNTech) | 85 326 300 | 84 023 380 | 169 349 680 |
| mRNA-1273 (Moderna) | 16 306 586 | 16 090 036 | 32 396 622 |
| ChAdOx1 nCoV-19 (Astra Zeneca) | 58 549 | 58 300 | 116 849 |
| Total | | | 201 863 151 |

**Table S2. Calculation of the average SARS-CoV-2 vaccination interval between the first and second dose.**

| | Interval between first and second dose [*a] | Number of second dose [*b] | Weighted average interval (days) |
|---|---|---|---|
| BNT162b2 (Pfizer-BioNTech) | 21 | 84 023 380 | |
| mRNA-1273 (Moderna) | 28 | 16 090 036 | |
| ChAdOx1 nCoV-19 (Astra Zeneca) | 40 | 58 300 | |
| Total | | 100 171 716 | 22.14 |

*a: Taken from the vaccine package insert.

*b: Information at 14 February 2022.

**Table S3. Characteristics of all-cause and myocarditis death in reference population.**

| | All-cause N (%) | | | Myocarditis N (%) | | |
|---|---|---|---|---|---|---|
| Year | 2017 | 2018 | 2019 | 2017 | 2018 | 2019 |
| Total | 1 337 761 (100) | 1 359 714 (100) | 1 378 395 (100) | 152 (100) | 166 (100) | 145 (100) |
| **Age (years)** | | | | | | |
| 10-19 | 1 598 (0.1) | 1 606 (0.1) | 1 603 (0.1) | 7 (4.6) | 6 (3.6) | 3 (2.1) |
| 20-29 | 4 302 (0.3) | 4 222 (0.3) | 4 097 (0.3) | 5 (3.3) | 13 (7.8) | 2 (1.4) |
| 30-39 | 8 004 (0.6) | 7 724 (0.6) | 7 455 (0.6) | 7 (4.6) | 12 (7.2) | 6 (4.1) |
| 40-49 | 22 838 (1.7) | 22 339 (1.6) | 21 837 (1.7) | 10 (6.6) | 15 (9.0) | 23 (15.9) |
| 50-59 | 46 591 (3.5) | 46 873 (3.4) | 46 935 (3.5) | 19 (12.5) | 20 (12.0) | 15 (10.3) |
| 60-69 | 137 354 (10.3) | 129 720 (9.5) | 121 635 (10.3) | 26 (17.1) | 39 (23.5) | 27 (18.6) |
| 70-79 | 264 972 (19.8) | 272 804 (20.1) | 281 734 (19.8) | 37 (24.3) | 30 (18.1) | 34 (23.4) |
| 80-89 | 496 298 (37.1) | 501 838 (36.9) | 501 343 (37.1) | 34 (22.4) | 24 (14.5) | 26 (17.9) |
| ≥ 90 | 355 335 (26.6) | 372 189 (27.4) | 391 272 (26.6) | 7 (4.6) | 7 (4.2) | 9 (17.9) |
| Unknown | 469 (0.0) | 399 (0.0) | 484 (0.0) | 0 (0) | 0 (0) | 0 (0) |
| **Sex** | | | | | | |
| Male | 689 264 (51.5) | 697 666 (51.3) | 705 983 (51.5) | 99 (65.1) | 91 (54.8) | 83 (57.2) |
| Female | 648 497 (48.5) | 662 048 (48.7) | 672 412 (48.5) | 53 (34.9) | 75 (45.2) | 62 (42.8) |
| Unknown | 0 (0) | 0 (0) | 0 (0) | 0 (0) | 0 (0) | 0 (0) |

**Table S4| Rate ratio for all-cause death by age by dose of vaccination.**

| | First dose | | | | | Second dose | | | | |
|---|---|---|---|---|---|---|---|---|---|---|
| Age (years) | Ob | Ex | Rate ratio | LL | UL | Ob | Ex | Rate ratio | LL | UL |
| 12-19 | 4 | 60 | 0.07 | 0.02 | 0.17 | 2 | 73 | 0.03 | 0.00 | 0.10 |
| 20-29 | 12 | 210 | 0.06 | 0.03 | 0.10 | 19 | 260 | 0.07 | 0.04 | 0.11 |
| 30-39 | 8 | 371 | 0.02 | 0.01 | 0.04 | 24 | 462 | 0.05 | 0.03 | 0.07 |
| 40-49 | 19 | 1 119 | 0.02 | 0.01 | 0.03 | 33 | 1 399 | 0.02 | 0.02 | 0.03 |
| 50-59 | 42 | 2 729 | 0.02 | 0.01 | 0.02 | 48 | 3 423 | 0.01 | 0.01 | 0.02 |
| 60-69 | 71 | 6 471 | 0.01 | 0.01 | 0.01 | 50 | 8 133 | 0.01 | 0.00 | 0.01 |
| 70-79 | 166 | 16 744 | 0.01 | 0.01 | 0.01 | 123 | 21 042 | 0.01 | 0.00 | 0.01 |
| 80-89 | 272 | 30 594 | 0.01 | 0.01 | 0.01 | 169 | 38 359 | 0.00 | 0.00 | 0.01 |
| ≥ 90 | 162 | 24 256 | 0.01 | 0.01 | 0.01 | 95 | 30 300 | 0.00 | 0.00 | 0.00 |
| Crude[*a] | 758 | 70 765 | 0.01 | 0.01 | 0.01 | 563 | 88 701 | 0.01 | 0.01 | 0.01 |
| SMR | | | 0.01 | 0.01 | 0.01 | | | 0.01 | 0.01 | 0.01 |

*a: Total all-cause deaths did not equal totals for each age group because they include persons of unknown age.

Abbreviations

Observed: Ob, Expected: Ex, Lower limit of 95% confidence interval: LL, Upper limit of 95% confidence interval: UL,

Standardized mortality ratio: SMR.

**Table S4(continued)| Rate ratio for all-cause death by age by dose of vaccination.**

| | Overall | | | | |
|---|---|---|---|---|---|
| Age (years) | Ob | Ex | Rate ratio | LL | UL |
| 12-19 | 7 | 133 | 0.05 | 0.02 | 0.11 |
| 20-29 | 31 | 470 | 0.07 | 0.04 | 0.09 |
| 30-39 | 34 | 833 | 0.04 | 0.03 | 0.06 |
| 40-49 | 54 | 2 518 | 0.02 | 0.02 | 0.03 |
| 50-59 | 93 | 6 151 | 0.01 | 0.01 | 0.02 |
| 60-69 | 126 | 14 604 | 0.01 | 0.01 | 0.01 |
| 70-79 | 302 | 37 786 | 0.01 | 0.01 | 0.01 |
| 80-89 | 458 | 68 953 | 0.01 | 0.01 | 0.01 |
| ≥ 90 | 267 | 54 555 | 0.00 | 0.00 | 0.01 |
| Crude[*a] | 1 374 | 159 466 | 0.01 | 0.01 | 0.01 |
| SMR | | | 0.01 | 0.01 | 0.01 |

*a: Total all-cause deaths did not equal totals for each age group because they include persons of unknown age.

Abbreviations

Observed: Ob, Expected: Ex, Lower limit of 95% confidence interval: LL, Upper limit of 95% confidence interval: UL,

Standardized mortality ratio: SMR.

**Table S5| Mortality odds ratio for myocarditis death by age.**

| Age (years) | Mortality odds ratio (MOR) | LL | UL |
|---|---|---|---|
| 12-19 | 0.06 | 0.00 | 210.82 |
| 20-29 | 67.51 | 12.12 | 246.65 |
| 30-39 | 240.16 | 80.28 | 632.36 |
| 40-49 | 82.07 | 15.83 | 267.82 |
| 50-59 | 28.25 | 0.69 | 168.08 |
| 60-69 | 103.03 | 20.58 | 317.58 |
| 70-79 | 164.45 | 58.49 | 374.68 |
| 80-89 | 277.05 | 107.47 | 600.76 |
| ≥ 90 | 367.11 | 41.74 | 1 497.67 |
| Crude | 209.82 | 146.06 | 301.41 |
| Pooled [*a] | 148.49 | 89.18 | 247.25 |

*a: Pooled mortality odds ratios were expressed based on random effect model.

Abbreviations

Observed: Ob, Expected: Ex, Lower limit of 95% confidence interval: LL,
Upper limit of 95% confidence interval: UL,
Standardized mortality ratio: SMR.

**Table S6 | Causal inference of increased risk of myocarditis mortality and SARS-CoV-2 vaccine use by applying modified US Surgeon General's criteria of causality referencing Hill's viewpoints.**

| # | US Surgeon General's Criteria | No of Hill's viewpoint | Comment | Status |
|---|---|---|---|---|
| 1 | Temporality of association | #4 | Very strong association; all events occurred after the vaccination. Moreover, onset of many death cases occurred within a short period after dose 1 or dose 2 of vaccine administration. Myocarditis symptoms leading to death occurred within 1 week in 50 % and within 2 weeks in 72 % of myocarditis death cases. | Y |
| 2 | Consistency of association | #2 | Similar risk levels on the incidence of myocarditis and/or pericarditis were reported from various countries[14-18], although they did not take healthy-vaccinee effect into account. | Y |
| 3 | Strength of association | #1 | 1) Strong association with high myocarditis mortality rate ratio (MMRR): Even without consideration of healthy-vaccinee effect, association measures for the risk of myocarditis death in the age of 30s were very strong (MMRR= 7.80). If healthy-vaccinee effect were taken into account, MMRR may be more than 7.0 for overall age which shows very strong association. Approximate 150 of pooled mortality odds ratio (MOR) also showed strong association. | Y |
| | | #5 | 2) Biological gradient (dose-response): Most of the epidemiological studies showed that the incidence rate of myocarditis or myopericarditis after the 2nd dose was higher than that after 1st dose. For example, according to the data by Mevorah et al[14] rate ratio of 2nd dose of BNT122b2 to 1st dose was 3.76 (95%CI: 2.40 to 5.90) for all and 8.40 (2.08 to 33.83) for male aged 16 to 19; By the other all 3 studies,[16-18] rate ratios of 2nd dose of mRNA-1273 to 1st dose were 5.06 or higher (lower limit of 95%CI were all 1.6 or higher). Our study showed that ratio of SMR after 2nd dose to that after 1st dose (1.36; 0.69 to 2.68) was not significant but is rather consistent with the tendency of the gradient for the incidence rate ratios of myocarditis from other studies. | |
| | | #3 | 3) Specificity: Myocarditis death is not a specific adverse reaction to SARS-CoV-2 vaccine and myocarditis death is caused not only SARS-CoV-2 vaccine but also by other vaccines and viruses or as an autoimmune disease. However, this is not an obstacle for satisfaction of this criterion "strength of association". | |

Y=criterion satisfied. X= criterion not satisfied.

**Table S6 (continued) | Causal inference of increased risk of myocarditis mortality and SARS-CoV-2 vaccine use by applying modified US Surgeon General's criteria of causality referencing Hill's viewpoints.**

| # | US Surgeon General's Criteria | No of Hill's viewpoint | Comment | Status |
|---|---|---|---|---|
| 4 | Coherence of association | #6 | 1) Biological plausibility: Some biological mechanisms were suggested such as increased interleukin-18 dependent immune responses to explain the association of myocarditis and vaccination.[59] | Y |
| | | #7 | 2) Coherence: There is no conflicting findings that deny the causality of myocarditis and SARS-CoV-2 vaccines. | |
| | | #8 | 3) Experiment: Randomized controlled trials did not report increased risk of myocarditis, but the population of the RCTs were not large enough to detect the risk. Real world use of vaccination may be considered as semi-experiment which was included in this viewpoint (#8) by Hill. Many studies on the semi-experiment reported the association almost without exception. | |
| | | #9 | 4) Analogy: Other vaccines (e.g., smallpox) have been reported to be associated with myocarditis or myopericarditis as one of the adverse reactions.[60] Three different products of SARS-CoV-2 vaccine (ChAdOx1, BNT122b2 and mRNA-1273) showed association of increased risk of myocarditis, although the highest risk was reported for mRNA-1273. | |

Y=criterion satisfied. X= criterion not satisfied.

59. Won T, Gilotra NA, Wood MK, et al. Increased Interleukin 18-Dependent Immune Responses Are Associated With Myopericarditis After COVID-19 mRNA Vaccination *Front Immunol* 2022; 13: 851620. doi: 10.3389/fimmu.2022.851620.

60. Decker MD, Garman PM, Hughes H, et al. Enhanced safety surveillance study of ACAM2000 smallpox vaccine among US military service members. *Vaccine* 2021; 39(39): 5541-47. doi: 10.1016/j.vaccine.2021.08.041.

| Vaccine type | Case No | Age | Sex | Last vaccinated date | Date of death | Number of vaccination | Comorbidities, histories leading to death | Cause of death diagnosed by physician | Corresponding MedDRA terms | Tests for the base of diagnosis | Causal assessment by physician | Causal assessment by experts |
|---|---|---|---|---|---|---|---|---|---|---|---|---|
| mRNA -1273 (Moderna) | 44 | 24 | Male | 14 August 2021 | 17 August 2021 | 2nd | Details unknown. The first vaccination was given on 17 July 2021. He developed a fever up to 38 °C, the day after the second vaccination. Two days after vaccination, the fever broke, but he complained of a headache. Three days after the vaccination he did not attend work and was found dead at home. | Acute myocarditis | Myocarditis | Autopsy (lymphocyte-dominated inflammatory cell infiltrate around myocardial arterioles. Diagnosis of acute myocarditis as cause of death, as there was necrosis of the myocardium, scattered fibrosis, mild lymphocytic infiltration of pericoronary adipose tissue and no other evidence of fatal endogenous lesion damage.) | Could not be assessed | Could not be assessed |

Available at: https://www.mhlw.go.jp/content/10601000/000961526.pdf
[in Japanese] (Accessed 13 December 2022)

**Fig S1. Examples of description in the Summary List of Deaths Following SARS-CoV-2 vaccination used by Japan's Ministry of Health, Labour and Welfare's Expert Committee on vaccination and adverse reactions.**

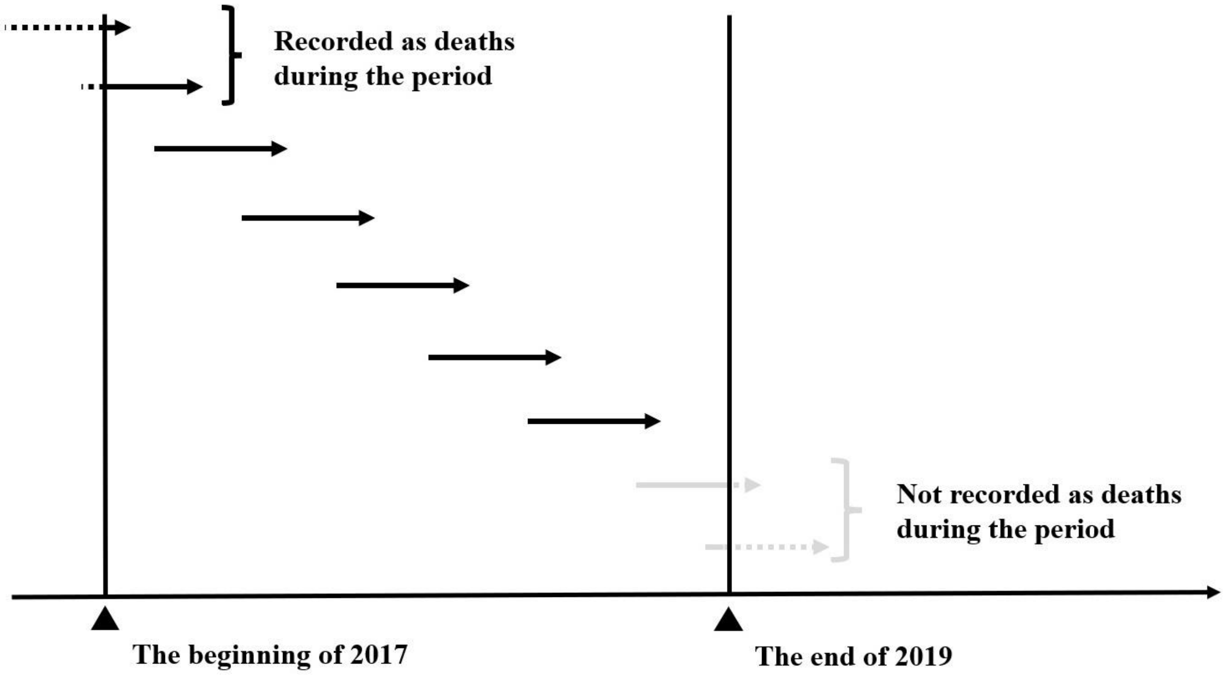


**Arrow lengths indicate time from onset to death.**

**Fig S2a. Method for counting deaths in the reference population.**

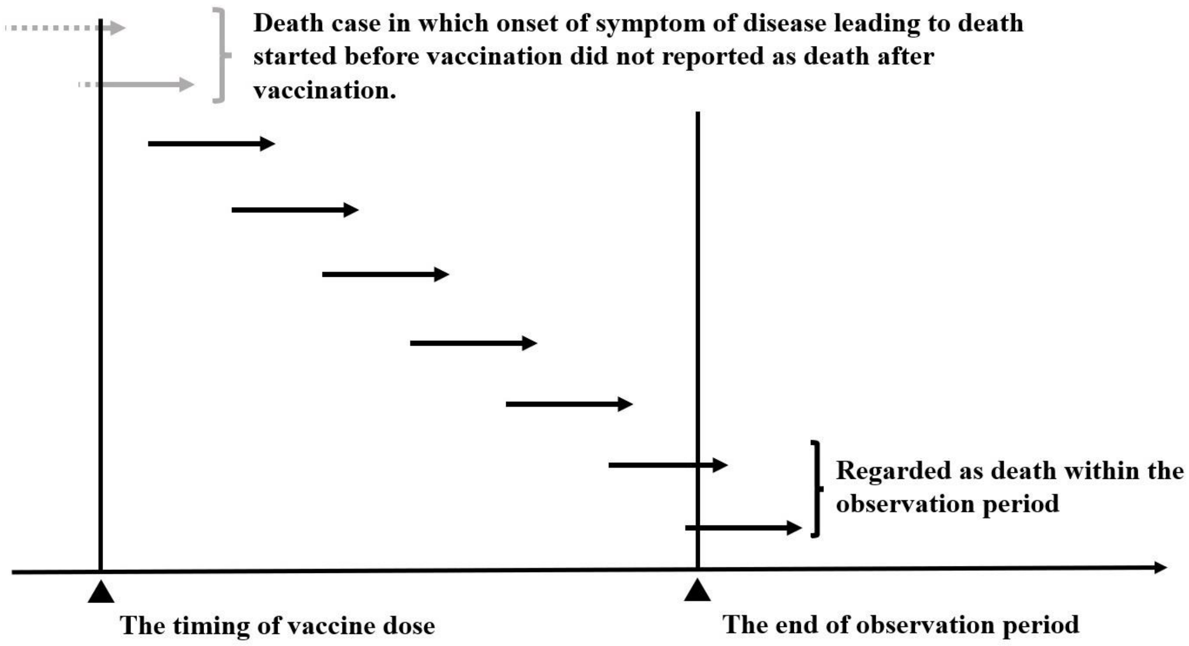


Arrow lengths indicate time from onset to death.

**Fig S2b. Method for counting deaths in the vaccinated population.**
